# Adding salt to foods and risk of incident infection

**DOI:** 10.64898/2026.08.26.26361489

**Authors:** Yu Qiao, Xinpei Chen, Yuyang Wang, Wei Zhao, Xizi Zheng, Xianghui Zhang, Guodong Niu, Yangfeng Wu, Yifang Yuan

## Abstract

**Background:** Excess sodium intake is a major contributor to the global burden of disease, but its role in infection susceptibility remains largely unexplored. Although sodium has been considered antimicrobial, high sodium intake may impair immune responses and host defense. We therefore examined whether habitual addition of salt to foods was associated with the long-term risk of incident infections.

**Methods:** We included 360,314 UK Biobank participants without prior hospital-treated infections. Frequency of adding salt to foods was self-reported at baseline. Incident infections were identified using ICD-10 codes from hospital and death records. Associations were assessed using multivariable Cox regression.

**Results:** Over a median follow-up of 14.3 years, 84 146 participants developed hospital-treated infections. Compared with those who never or rarely added salt, participants who sometimes, usually, and always added salt had progressively higher risks of incident infections (adjusted hazard ratios 1.04 [95% CI 1.03–1.06], 1.08 [1.06–1.11], and 1.29 [1.26–1.33], respectively; p for trend <0.001). The association remained robust across models and broadly consistent across pathogen types and infection sites. The association appeared stronger among participants with normal weight (P for interaction <0.001).

**Conclusions:** Habitual addition of salt to foods was associated with a dose-dependent higher risk of hospital-treated infections in this large prospective cohort. These findings extend the potential health relevance of excess sodium intake beyond cardiometabolic disease and suggest that lower habitual salt intake may have implications for infection risk. Further studies are needed to replicate these findings and clarify the underlying immunological mechanisms.

**Funding:** The study was supported by the National Natural Science Foundation of China, Ministry of Science and Technology of China (No.82404360).

## Introduction

Excessive salt intake has emerged as a global public health challenge, attracting significant attention in recent years. Extensive evidence has established a robust link between dietary sodium intake and the risk of hypertension and cardiovascular disease^1,2^. Trials have demonstrated that reducing sodium intake leads to significant reductions in all-cause mortality^3^, primarily driven by decreased cardiovascular risk. In 2021, a diet high in sodium was responsible for 41·3 million DALYs and 1·86 million deaths^4^. It was the second-leading dietary risk factor for attributable DALYs. The World Health Organization (WHO) recommends that adults consume less than 5 grams of salt per day (equivalent to 2000 mg of sodium) and list sodium reduction as one of the “best-buys” to prevent noncommunicable diseases^5^.

Despite the well-established risks of excessive sodium intake, the physiological role of sodium presents a more complex story. Sodium may be an ancient way to ward off microbes. Historically, high-sodium has been vital for food preservation, leveraging its antimicrobial properties to inhibit spoilage. Prior studies^6^ suggested that a high-salt diet may increase the accumulation of skin Na+ storage and improve defenses against certain infections, such as cutaneous leishmaniasis. However, more recent studies indicate that excessive sodium intake could weaken immune responses and increase susceptibility to infections^7^. Most of these findings come from animal models, with conflicting results and limited evidence on human populations. It is unclear whether the restriction of salt intake would lead to an unexpected side effect: an increased risk of infection.

Self-reported data on "adding salt to food" at the table offers a practical proxy for habitual salt consumption. It correlates with 24-hour sodium excretion^8,9^ and may reflect long-term behaviors with minimal influence from daily variations. Additionally, this simple measure may be easily translatable into public health messaging. In this study, we used self-reported salt-adding behavior as a marker of dietary sodium intake to explore its association with incident infection in the UK Biobank. This investigation may provide new insights into the broader health implications of dietary sodium consumption and inform future public health recommendations.

## Methods

### Study population

The UK Biobank is a prospective cohort study, which recruited more than 0.5 million community-dwelling adults, aged 37 to 73, at 22 assessment centers across the UK from 2006 to 2010.^10^ The detailed study design has been described elsewhere.^11^ All participants provided written informed consent. The UK Biobank received ethical approval from the North West Multi-Centre Research Ethics Committee and the present study was approved by the UK Biobank (application ID 179610).

In this study, we pooled data from 502 154 UK Biobank participants. We excluded individuals with a history of hospital-treated infections (n = 50 998), those with incomplete data on the frequency of adding salt to foods at baseline (n = 909), and those with incomplete covariate data (n = 89 933). Finally, a total of 360 314 participants were included in the main analysis, as presented in **Figure 1**.

**Figure 1.**
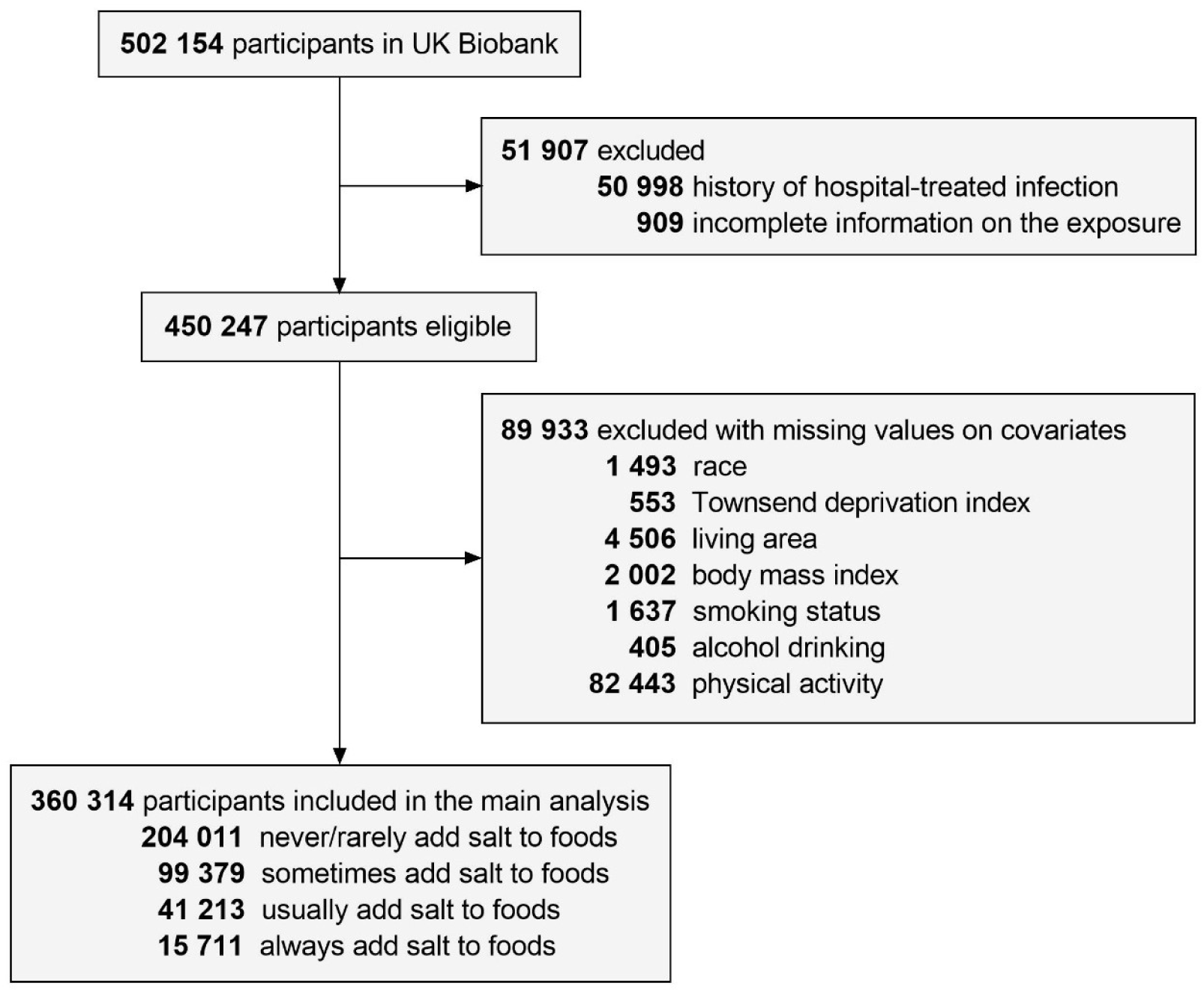
Flow chart of the present study

### Exposure assessment

The exposure of the study was self-reported frequency of adding salt to foods, which was measured with a touchscreen questionnaire at the assessment center (2006-2010). Participants were asked, “Do you add salt to your foods? (Do not include salt used in cooking),” and they chose 1 answer from the following 5 options: 1) never/rarely; 2) sometimes; 3) usually; 4) always; or 5) prefer not to answer. Those who preferred not to answer were considered to have missing information and were excluded from the analysis. The exposure was treated as a four-level categorical variable and participants who never or rarely added salt to foods were the reference.

### Identification of outcome

The primary outcome of the present study was the occurrence of any infection after the baseline. We extracted the diagnoses of infectious diseases from inpatient hospital discharge information and death registries using the International Classification of Disease 10th edition (ICD-10) codes^12^. The secondary outcomes were defined based on different types (bacteria, virus, fungus, and parasitic infection)^12^ and locations (respiratory tract, urinal tract, digestive tract, skin infection, and sepsis)^13^ of the infections according to ICD-10 codes (**Table S1**). The follow-up time was calculated from the date of attending the assessment center to the diagnoses of the outcomes, death, or the censoring date (31 October 2023), whichever came first.

### Covariates

A range of covariates were also assessed, including age, sex, race, Townsend deprivation index (TDI), living area, body mass index (BMI), diet quality, physical activity, smoking status, alcohol drinking, C-reactive protein (CRP) level, urinary potassium, non-steroidal anti-inflammatory drug (NSAID) or immunosuppressive agent (ISA) medication, prevalent hypertension, diabetes mellitus (DM), cardiovascular disease (CVD), chronic kidney disease (CKD), and cancer. The demographical and lifestyle factors were obtained through standard touch-screen questionnaires. Race was categorized as White and non-White (including Black, Asian, mixed, and others). The TDI^14^ is an area-specific composite measurement of material deprivation based on the participants’ postcode, which was calculated with 4 domains (non-car ownership, non-home ownership, unemployment, and overcrowding). Diet quality (healthy vs unhealthy) was estimated based on information of 7 food groups’ intake: fruits, vegetables, fish, unprocessed red meats, processed meats, whole grains, and refined grains^15^. Regular physical activity was defined as ≥ 150 minutes moderate activity per week or ≥ 75 minutes vigorous activity per week or their equivalent combination^16^. Moderate alcohol drinking was defined as pure alcohol intake > 0 and ≤ 14g/day for women, and > 0 and ≤ 28g/day for men. CRP and spot urinary potassium were measured on the blood and urinary samples collected in the assessment center, respectively. Self-reported information on using of NSAID or ISA was obtained in the verbal interview process in the assessment center^17^, with the corresponding drug codes listed in **table S2**. Prevalent chronic conditions were acquired through self-reported data, medication history, hospital inpatient data, and relevant measurements at the assessment center. The details of covariate definition and related data fields in UK Biobank are presented in **table S3**. **Statistical analysis**

We used the complete case dataset for the main analysis **(Table S4**). Cox proportional hazard regression models were employed to evaluate the association of the frequency of adding salt to foods (as a categorical variable) with the primary and secondary outcomes. P for trend was derived by modeling the frequency of adding salt as a continuous variable in the Cox regression model. The proportional hazards assumptions were not violated by Schoenfeld residuals method. We adjusted for covariates including age, sex, race, TDI, BMI, current smoking, moderate alcohol drinking, and regular physical activity in the main models. To test the robustness of the results, we performed the following sensitivity analyses: (1) further adjusting for other potential confounders based on the main models: history of hypertension, urinary potassium, diet quality, elevated CRP level, and NSAID/ISA medication; (2) excluding participants with prevalent DM, CVD, CKD or cancer at baseline; (3) excluding participants with infection within the first 2 years of follow-up; (4) excluding participants with self-reported dietary change over the past 5 years.

We performed subgroup analyses of the primary outcome (any infection) by the following potential risk factors: age (< 60 or ≥ 60 years), sex (men or women), race (Whites or non-Whites), TDI (quintile 1, quintiles 2–4, or quintile 5), BMI (normal weight, or overweight/obesity), smoking status (current smoker or not), alcohol drinking (moderate drinking or not), regular physical activity (yes or no), diet quality (healthy or unhealthy), history of hypertension (yes or no), spot urinary potassium (quintile 1, quintiles 2–4, or quintile 5), and CRP level (< 2 or ≥ 2 mg/L). To evaluate the interactions between the frequency of adding salt to foods and these factors, we added the interaction terms to the Cox models. The overall p values for interaction were calculated with likelihood ratio test by comparing models with or without the interaction term. All covariates adjusted in these models were consistent with those in the main models except the stratifying variable. All statistical analyses were performed with R version 4.2.3 (R Foundation) using the “survival” packages. A two-sided P-value of < 0.05 was considered statistically significant.

## Results

### Baseline characteristics

A total of 360 314 participants (mean age 56.2 ± 8.1 years, female sex 52.9%) were included in the main analysis, in which 204 011 (56.6%), 99 379 (27.6%), 41 213 (11.4%), and 15 711 (4.4%) participants reported that they never/rarely, sometimes, usually, and always added salt to foods, respectively. Compared with participants with lower frequency of adding salt to foods, participants with higher frequency were more likely to be men and non-white race, to have lower socioeconomic status (higher TDI), unhealthier lifestyles (higher prevalence of obesity and current smoking and lower prevalence of moderate drinking, healthy diet, and regular physical activity), and higher inflammatory status (higher CRP level), and had higher prevalence of DM, CVD, CKD, and NSAID or ISA medication. The baseline characteristics of the study population according to the frequency of adding salt to foods are shown in **Table 1**.

**Table 1.** Baseline characteristics of the study population.

| Variables | Total | Frequency of adding salt to foods |  |  |  | P value |
| --- | --- | --- | --- | --- | --- | --- |
|  |  | Never/rarely | Sometimes | Usually | Always |  |
| Number of participants | 360 314 | 204 011 | 99 379 | 41 213 | 15 711 |  |
| Age, years, mean (SD) | 56.2 (8.1) | 56.2 (8.1) | 56.1 (8.1) | 56.7 (8.0) | 55.8 (8.2) | 0.004 |
| Female sex (%) | 52.9 | 54.4 | 52.5 | 47.6 | 50.1 | <0.001 |
| White race (%) | 95.3 | 96.3 | 94.6 | 94.5 | 89.7 | <0.001 |
| Townsend deprivation index (%) |  |  |  |  |  | < 0.001 |
| Low (Q1) | 21.2 | 22.2 | 20.6 | 19.7 | 15.3 |  |
| Intermediate (Q2-Q4) | 61.0 | 61.5 | 60.8 | 60.4 | 56.0 |  |
| High (Q5) | 17.9 | 16.3 | 18.6 | 19.9 | 28.7 |  |
| Living in urban (%) | 85.5 | 85.2 | 85.6 | 85.7 | 88.1 | < 0.001 |
| Body mass index (%) |  |  |  |  |  | < 0.001 |
| Normal weight | 34.3 | 36.5 | 32.3 | 29.9 | 29.3 |  |
| Overweight | 43.0 | 42.4 | 43.7 | 44.4 | 42.6 |  |
| Obesity | 22.7 | 21.1 | 24.0 | 25.7 | 28.2 |  |
| Current smoker (%) | 9.7 | 7.5 | 10.5 | 14.2 | 21.8 | < 0.001 |
| Moderate drinking (%) | 65.0 | 66.0 | 65.7 | 62.1 | 54.6 | < 0.001 |
| Regular physical activity (%) | 61.3 | 62.0 | 61.4 | 59.6 | 56.7 | < 0.001 |
| Healthy diet (%) | 70.2 | 74.4 | 67.6 | 60.4 | 51.4 | < 0.001 |
| Changing diet during the past five years (%) | 38.1 | 38.7 | 38.2 | 36.7 | 35.5 | < 0.001 |
| High-sensitive C-reactive protein, mg/L, median (IQR) | 1.25 (0.62 - 2.57) | 1.18 (0.59 - 2.43) | 1.30 (0.65 - 2.64) | 1.38 (0.69 - 2.82) | 1.57 (0.77 - 3.28) | < 0.001 |
| Urinary potassium, mmol/L, median (IQR) | 57.3 (37.2 - 83.6) | 56.2 (36.5 - 82.4) | 58.1 (37.8 - 84.5) | 59.6 (38.9 - 86.1) | 59.4 (38.6 - 86.0) | < 0.001 |
| Hypertension (%) | 76.8 | 77.1 | 76.3 | 77.0 | 76.0 | <0.001 |
| Diabetes mellitus (%) | 6.1 | 6.0 | 6.2 | 6.3 | 6.6 | <0.001 |
| Cardiovascular disease (%) | 6.7 | 6.7 | 6.3 | 7.1 | 7.8 | <0.001 |
| Chronic kidney disease (%) | 5.8 | 5.8 | 5.7 | 5.8 | 6.4 | 0.003 |
| Cancer (%) | 4.2 | 4.2 | 4.2 | 4.4 | 4.2 | 0.145 |
| NSAID or ISA medication (%) | 9.8 | 8.9 | 10.4 | 11.5 | 12.8 | <0.001 |

IQR, interquartile range; ISA, immunosuppressive agent; NSAID, non-steroidal anti-inflammatory drug

### Adding salt and infection

During a median follow-up of 14.3 (IQR 13.3 – 15.2) years (4 633 264 person-years at risk), 84 146 participants (23.4%) were observed to have incident infectious disease, in which 69 404(19.3%), 8 926(2.5%), 4 961(1.4%), and 441(0.1%) participants had bacterial, viral, fungal, and parasitic infection, respectively, and 32 840(9.1%), 17 947(5.0%), 8 149(2.3%), 11 019(3.1%), and 10 687(3.0%) participants had respiratory tract, urinary tract, digestive tract, skin infection and sepsis, respectively.

After adjustment for age, sex, race, TDI, BMI, smoking, alcohol drinking, and physical activity, the hazard of any infection significantly increased with increasing frequency of adding salt to foods. The adjusted HRs were 1 (reference), 1.04 (1.03 - 1.06), 1.08 (1.06 - 1.11), and 1.29 (1.26 - 1.33) across groups, respectively (P for trend < 0.001) (**Table 2**). These results did not change appreciably after further adjusting for hypertension, spot urinary potassium, diet quality, CRP, or NSAID/ISA medication, or excluding participants with prevalent DM, CVD, CKD or cancer, or outcomes occurring within the first 2 years of follow-up, or those with dietary change over the past 5 years. (**Table 2**)

**Table 2.** Associations of frequency of adding salt to foods with any hospital-treated infection.

| Any hospital-treated infection | Never/rarely | Sometimes | Usually | Always | P for trend |
| --- | --- | --- | --- | --- | --- |
| Cases/total | 45264/204011 | 23486/99379 | 10612/41213 | 4784/15711 |  |
| Incidence per 100 000 person-years | 1549.4 | 1652.4 | 1810.6 | 2159.4 |  |
| Primary analysis <sup>a</sup> | 1 (Reference) | 1.04 (1.03 - 1.06) | 1.08 (1.06 - 1.11) | 1.29 (1.26 - 1.33) | < 0.001 |
| Sensitivity analyses |  |  |  |  |  |
| Maximal adjusted model <sup>b</sup> | 1 (Reference) | 1.03 (1.01 - 1.05) | 1.05 (1.02 - 1.08) | 1.25 (1.20 - 1.31) | < 0.001 |
| Excluding participants with DM,<br>CVD, CKD or cancer at baseline or<br>during follow-up | 1 (Reference) | 1.00 (0.93 - 1.08) | 1.17 (1.05 - 1.29) | 1.56 (1.36 - 1.79) | < 0.001 |
| Excluding participants with<br>infections within the first 2 years of<br>follow-up | 1 (Reference) | 1.04 (1.03 - 1.06) | 1.08 (1.06 - 1.11) | 1.29 (1.25 - 1.33) | < 0.001 |
| Excluding participants with dietary<br>change over the past 5 years | 1 (Reference) | 1.04 (1.02 - 1.06) | 1.09 (1.06 - 1.12) | 1.35 (1.30 - 1.40) | < 0.001 |

<sup>a</sup> adjusted for age, sex, race, Townsend deprivation index, living area, body mass index, smoking status, alcohol drinking, and physical activity.
<sup>b</sup> additionally adjusted for history of hypertension, urinary potassium, diet quality, elevated C-reactive protein level, and non-steroidal anti-inflammatory drug/immunosuppressive agent medication.
CKD, chronic kidney disease; CVD, cardiovascular disease; DM, diabetes mellitus.

Furthermore, a higher frequency of adding salt to foods was associated with a dose-dependent increase in the risk of different infectious types and locations (all P for trend < 0.001), with the point estimated of adjusted HRs ranging from 1.22 to 1.64 for those who always added salt compared to those who never or rarely did, except for the association with parasitic infections, in which the adjusted HR for participants who always added salt to foods was 1.64 [1.15–2.35], while the P for trend did not reach the statistical significance (P = 0.069) (Table 3). The results were robust in the sensitivity analyses. (**Table S5 and S6**)

**Table 3.** Associations of frequency of adding salt to foods with secondary outcomes.

| Variables | Never/rarely | Sometimes | Usually | Always | P for trend |
| --- | --- | --- | --- | --- | --- |
| **By infectious type** |  |  |  |  |  |
| **Bacterial infection** |  |  |  |  |  |
| Cases/total | 37168/204011 | 19442/99379 | 8779/41213 | 4015/15711 |  |
| Incidence per 100 000 person-years | 1272.3 | 1367.8 | 1497.8 | 1812.3 |  |
| Adjusted HR <sup>a</sup> | 1 (Reference) | 1.05 (1.03 - 1.07) | 1.08 (1.05 - 1.11) | 1.30 (1.26 - 1.34) | < 0.001 |
| **Viral infection** |  |  |  |  |  |
| Cases/total | 4793/204011 | 2502/99379 | 1099/41213 | 532/15711 |  |
| Incidence per 100 000 person-years | 164.1 | 176.0 | 187.5 | 240.1 |  |
| Adjusted HR <sup>a</sup> | 1 (Reference) | 1.03 (0.98 - 1.08) | 1.04 (0.97 - 1.11) | 1.25 (1.14 - 1.37) | < 0.001 |
| **Fungal infection** |  |  |  |  |  |
| Cases/total | 2555/204011 | 1442/99379 | 647/41213 | 317/15711 |  |
| Incidence per 100 000 person-years | 87.5 | 101.5 | 110.4 | 143.1 |  |
| Adjusted HR <sup>a</sup> | 1 (Reference) | 1.13 (1.05 - 1.20) | 1.13 (1.04 - 1.23) | 1.41 (1.25 - 1.59) | < 0.001 |
| **Parasitic infection** |  |  |  |  |  |
| Cases/total | 230/204011 | 123/99379 | 52/41213 | 36/15711 |  |
| Incidence per 100 000 person-years | 7.9 | 8.7 | 8.9 | 16.2 |  |
| Adjusted HR <sup>a</sup> | 1 (Reference) | 1.03 (0.83 - 1.28) | 1.01 (0.75 - 1.36) | 1.64 (1.15 - 2.35) | 0.069 |
| **By infectious location** |  |  |  |  |  |
| **Respiratory tract infection** |  |  |  |  |  |
| Cases/total | 17335/204011 | 9191/99379 | 4300/41213 | 2014/15711 |  |
| Incidence per 100 000 person-years | 593.4 | 646.6 | 733.6 | 909.1 |  |
| Adjusted HR <sup>a</sup> | 1 (Reference) | 1.06 (1.03 - 1.08) | 1.10 (1.06 - 1.13) | 1.34 (1.28 - 1.40) | < 0.001 |
| **Urinary tract infection** |  |  |  |  |  |
| Cases/total | 9669/204011 | 5046/99379 | 2213/41213 | 1019/15711 |  |
| Incidence per 100 000 person-years | 331.0 | 355.0 | 377.6 | 460.0 |  |
| Adjusted HR <sup>a</sup> | 1 (Reference) | 1.04 (1.00 - 1.07) | 1.02 (0.98 - 1.07) | 1.26 (1.18 - 1.35) | < 0.001 |
| **Digestive tract infection** |  |  |  |  |  |
| Cases/total | 4375/204011 | 2289/99379 | 1028/41213 | 457/15711 |  |
| Incidence per 100 000 person-years | 149.8 | 161.0 | 175.4 | 206.3 |  |
| Adjusted HR <sup>a</sup> | 1 (Reference) | 1.04 (0.99 - 1.09) | 1.08 (1.01 - 1.16) | 1.22 (1.11 - 1.35) | < 0.001 |
| **Skin infection** |  |  |  |  |  |
| Cases/total | 5809/204011 | 3141/99379 | 1430/41213 | 639/15711 |  |
| Incidence per 100 000 person-years | 198.8 | 221.0 | 244.0 | 288.4 |  |
| Adjusted HR <sup>a</sup> | 1 (Reference) | 1.06 (1.01 - 1.10) | 1.08 (1.02 - 1.14) | 1.23 (1.13 - 1.33) | < 0.001 |
| **Sepsis** |  |  |  |  |  |
| Cases/total | 5657/204011 | 2931/99379 | 1416/41213 | 683/15711 |  |
| Incidence per 100 000 person-years | 193.6 | 206.2 | 241.6 | 308.3 |  |
| Adjusted HR <sup>a</sup> | 1 (Reference) | 1.02 (0.98 - 1.07) | 1.08 (1.02 - 1.15) | 1.38 (1.27 - 1.49) | < 0.001 |

<sup>a</sup> the main model: adjusted for age, sex, race, Townsend deprivation index, living area, body mass index, smoking status, alcohol drinking, and physical activity.

### Subgroup analyses

Subgroup analysis revealed that the association between adding salt to foods and infection risk was generally consistent across different subgroups, except for a stronger association observed in individuals with normal weight compared to those with overweight or obesity (P for interaction < 0.001) (**Figure 2 and Table S7**).

**Figure 2.**
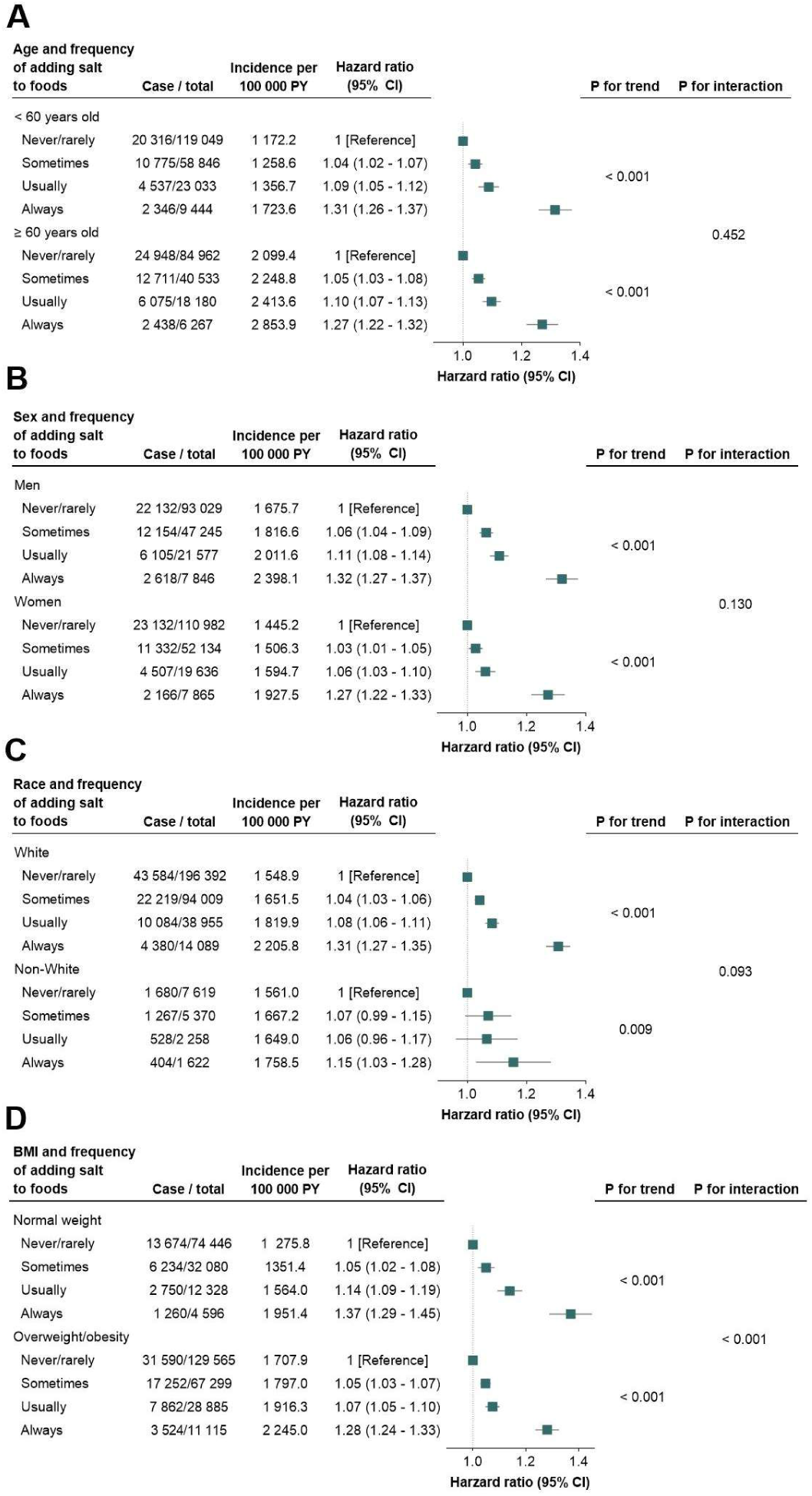
Interaction between age, sex, race, or body mass index and the association of adding salt to foods with risk of any infections. Multiplicative interaction was evaluated with likelihood ratio test by comparing models with or without the interaction term. All covariates adjusted in these models were consistent with those in the main models except the stratifying variable. BMI, body mass index; CI. confidence interval; PY, person-year.

## Discussion

In this cohort study of UK Biobank participants, we observed that a higher self-reported frequency of adding salt to foods was significantly associated with an increased risk of incident infections. Specifically, participants who always added salt to food at the table generally exhibited a 30-50% higher risk of developing new-onset infections compared to those who never or rarely added salt, after adjusting for SES, lifestyle factors, and other traditional risk factors of infection. Notably, this association appeared consistent across different types of pathogens and infection locations. These findings highlight the potential significance of maintaining a habitually lower salt intake as a preventive strategy for reducing infection risk. Public health efforts aimed at reducing salt consumption may contribute not only to cardiovascular benefits but also to enhanced immune resilience against infections.

To our knowledge, this is the first population study to systematically examine the association between sodium intake and incident infection. Our findings shed light on this underexplored area and emphasize the need for further research to clarify the effects of sodium on immunity and infection risk. Numerous studies have provided compelling evidence on the health impact of excessive sodium intake. Robust data indicate that high sodium intake is associated with increased CV risk^18^, primarily through its dose-response linear relationship with elevated blood pressure^19^. Furthermore, clinical trials showed that sodium reduction decreased all-cause mortality, largely attributable to its CV benefits^3^. The impact of sodium intake on non-cardiovascular outcomes such as stomach cancer^8^, osteoporosis^20^, kidney stones^21^, and atopic dermatitis^22^, remains exploratory and require validation with high-quality population studies and trials. The relationship between sodium intake and immune system as well as infection is even more uncertain. Some animal studies^6^ suggest we might “pickle” our cells to ward off the microbes by boosting macrophage activation and promoting cutaneous immunity. More recent studies propose that high sodium diets may impair immune function^7,23^, or with the effects varying by tissue type, immune cell involvement, and disease context^24^. These findings are largely derived from animal models or small human experiment and lack consistency. The present study offers novel population-based insights into the potential relationship between sodium intake and infection risk.

There are possible mechanisms that could help to explain the link between habitual high salt intake and increased risk of infection. High sodium intake increased glucocorticoid levels^24^, which can suppress immunity such as inhibiting neutrophil-mediated phagocytosis and thus impairing the clearance of pathogens. A one-week high-salt challenge in ten healthy volunteers^7^ resulted in hyperglucocorticoidism and reduced neutrophil-mediated antibacterial function. In addition to the upregulates of glucocorticoid, a downregulates of RAAS hormones may also weaken the innate immune system. For adaptive immune system, increasing NaCl may impair regulatory T cell (Treg) function in association with heightened IFN-γ (interferon-γ) secretion both in vivo and in vitro^25^. In addition, high dietary sodium intake can alter the gut microbiota and affect gut immune homeostasis^26^, which may have consequences for certain inflammatory process. High sodium intake may also exert localized effects, particularly on the kidney^7^. In the present study, the association between habitual salt addition and infection risk was consistent across various pathogens and infection sites. This suggests that a high-salt diet may subtly yet meaningfully disrupt the foundational “soil” of the immune system, thus impairing the body’s capacity to defend against infections. We conducted a mediation analysis (data not shown), which demonstrated that CRP accounted for a modest 4.3% of the association, further supporting the notion that the effect of sodium on infection risk is systemic in nature rather than driven by a specific immune cell type or singular biological pathway. Overall, these findings highlight the complex but notable impact of high sodium intake on immune system and infection risk.

The subgroup analysis suggests that the association between added salt intake and incident infection was generally consistent, except that stronger association was observed among individuals with normal weight. Notably, the attenuation of salt’s impact with increasing BMI aligns with prior findings on premature mortality^9^, cardiovascular disease^27^, and chronic kidney disease^28^. One plausible explanation is that obesity-related chronic inflammation and pre-existing immune activation ^29^ may mask the additional immunomodulatory effects of salt, making the additional risk less discernible in individuals with higher BMI. Another consideration is that the normal weight group may include a small proportion of individuals with underweight and potential malnutrition, who may be more vulnerable to infections due to inadequate physiological and nutritional reserves^30^. While these subgroup findings provide valuable insights, they should be interpreted with caution and further research is needed to validate these observations and elucidate the underlying mechanisms.The findings of the present study do not totally deny the message from prior research that high salt intake may play a role in combating microbes. Instead, they highlight the distinction between local and systemic effects. In the context of food preservation, salt inhibits microbial growth, reducing the risk of contamination. However, this study focuses on the systemic impact of dietary salt intake on the “soil” of the immune system and its potential to increase infection susceptibility. The overall potential detrimental effect of a high-salt does not deny the possibility of its localized antimicrobial benefits. For example, while increased sodium storage in skin may activate macrophages to promote antimicrobial activity^6^, direct sodium exposure such as topical application, rather than dietary intake via the gastrointestinal tract, may more efficiently achieve the necessary local concentrations to exert antimicrobial effect.

The strengths of this study lie in its prospective design, large sample size, comprehensive covariate data collection, and consistent findings across sensitivity analyses. However, several limitations should be acknowledged. First, we did not employ the gold standard multiple measurements of 24-hour urinary sodium excretion to assess dietary sodium intake, as this information was unavailable in the UK Biobank. However, the self-reported frequency of adding salt to foods can serve as a surrogate marker for long-term sodium intake. This measure has been shown to exhibit a graded association with estimated 24-hour sodium excretion^8,9^, and reflects not only short-term sodium intake but also a preference for salty taste in foods and habitual salt consumption, with less susceptibility to daily variation^8,27,28^. In addition, this simple measure may be easily incorporated into public health messaging. Second, our study population was based in the United Kingdom, where sodium reduction policies have been relatively successful, leading to lower overall sodium intake compared to other countries such as the United States and China. T In addition, cultural differences in dietary habits, particularly the limited use of table salt in other cultures like Chinese cuisine, may affect the interpretation of adding salt to foods, despite the underlying biological mechanisms remaining consistent. Third, while the study did not exhaustively include all types of infections, we tried to cover all clinically meaningful categories commonly used in medical practice. Finally, although residual confounding cannot be ruled out, the results from sensitivity analyses were stable, supporting the robustness of our findings.

## Conclusion

In conclusion, our study demonstrates that a higher self-reported frequency of adding salt to foods is significantly associated with an increased risk of incident infections. This association is broadly consistent across various pathogen types and infection sites. These findings underscore the potential importance of habitual lower salt intake as a strategy for infections prevention. Further independent studies are warranted to confirm our findings and its implications.

## Data Availability

This research was conducted using the UK Biobank(application ID: 179610). The data are available to bonafide researchers through UK Biobank upon application. Analytic methods and study materials are available from the corresponding author upon reasonable request

## ACKNOWLEDGEMENTS

The authors thank all investigators for their proactive participation in the study. We are also grateful to Prof Li Weng at the Peking Union Medical College Hospital for his advice on study design.

## Contributors

YQ and YY designed the study with advices from WZ and YW. YQ analyzed and verified the data analysis. YY and YW helped on data analysis and interpretation. YY, YQ and YW wrote the first draft with all co-authors participating in the subsequent reviews and revisions. YY, YW and GN had full access to all of the data in the study and take responsibility for the integrity of the data and the accuracy of the data analysis.

